# Patient-collected tongue, nasal, and mid-turbinate swabs for SARS-CoV-2 yield equivalent sensitivity to health care worker collected nasopharyngeal swabs

**DOI:** 10.1101/2020.04.01.20050005

**Authors:** YP Tu, R Jennings, B Hart, GA Cangelosi, RC Wood, K Wehber, P Verma, D Vojta, EM Berke

## Abstract

**Background:** Current testing for SARS-CoV-2 requires health care workers to collect a nasopharyngeal (NP) sample from a patient. NP sampling requires the use of personal protective equipment that are in limited supply, is uncomfortable for the patient, and reduces clinical efficiency. This study explored the equivalency of patient-collected tongue, anterior nares (nasal), and mid-turbinate (MT) samples to health care worker-collected NP samples for detecting SARS-CoV-2.

**Methods:** Patients presenting to five urgent care facilities with symptoms indicative of an upper respiratory infection provided self-collected samples from three anatomic sites along with a health care worker-collected NP sample. Using NP as the comparator, sensitivities and one-sided 95% confidence intervals for the tongue, nasal, and MT samples for detection of SARS-CoV-2 were calculated.

**Results:** The sensitivity for detecting SARS-CoV-2 in patient-collected tongue, nasal, and mid-turbinate samples was 89.8% (95% CI: 80.2 -100.0), 94.0 (95% CI: 84.6-100.0) and 96.2 (95% CI: 87.7-100.0), respectively. Among samples yielding positive results, cycle threshold (Ct) values (a measure of viral load) had correlation coefficients of 0.48, 0.78, and 0.86 between the NP samples and the tongue, nasal, and MT samples, respectively.

**Conclusions:** Patient-collected nasal and MT samples demonstrated high sensitivity for SARS-CoV-2 detection using health care worker-collected NP samples as the comparator. Among patients testing positive with NP samples, nasal and MT Ct values demonstrated high correlations with those Ct values of the NP samples. Patient-collected nasal or MT sampling may improve efficiency for COVID-19 testing while reducing the risk of exposure of the health workforce.

## Introduction

The early medical response to the COVID-19 pandemic in the United States has been highlighted by limitations in the availability of testing among symptomatic people. By the time the total number of confirmed cases in the United States reached 33,404 on March 23, 2020 with 400 deaths^1^, public health officials in areas with high proportion of cases recommended against ambulatory testing in favor of higher risk individuals^2,3^. In vitro diagnostic testing in the face of epidemic spread to provide both clinical care and inform public health efforts is well established^4^. Current guidelines for testing of people with suspected COVID-19 require a swab of the oropharynx (OP) or nasopharynx (NP) to extract and amplify any viral RNA by real-time reverse transcription-polymerase chain reaction (rRT-PCR)^5^. Transmission of SARS-CoV-2 to health care workers has been described extensively^6,7^. The use of personal protective equipment (PPE) by health care workers obtaining testing samples is critical to reduce transmission, but there are shortages of such equipment in many hospitals^8^.

For other virus-mediated upper respiratory infections, such as influenza, viral material can be detected from swabs of the lower nares and mid-turbinate region^9,10,11^. Experience with respiratory pathogens such as tuberculosis have also shown that samples obtained from tongue-swabs have sufficient accuracy for diagnosis^12,13^. In these other clinical experiences, obtaining a tongue, nasal, or mid-turbinate (MT) sample is faster, better tolerated, and causes less potential for sneezing, coughing and gagging, than an NP swab. Additional recent evidence supports the validity of non-NP samples for SARS-CoV-2 detection^14,15^.

We investigate whether self-collected tongue, nasal, or MT samples from symptomatic people with suspected COVID-19 are equivalent to health care worker-collected NP samples for detecting SARS-CoV-2.

## Methods

### Population and Sample Collection

People seen in any one of five ambulatory clinics in the Puget Sound region with symptoms indicative of upper respiratory infection between the dates March 16 and March 21 were eligible for participation. We enrolled all people who were willing and able to participate in the self-collection of all three anatomic sites: tongue, nasal and MT and health care worker-collection from the NP. Inclusion criteria included evidence of symptoms suggestive of an upper respiratory illness (subjective and objective fevers, cough, sore throat, fevers, myalgia, or rhinorrhea, indicating higher risk of COVID-19 in this community) and the ability to consent and agree to participate in the study. People who were not able to demonstrate understanding of the study, not willing to commit to having all four samples collected, had a history of nosebleed in the past 24 hours, nasal surgery in the past two weeks, chemotherapy treatment with documented low platelet and low white blood cell counts, or acute facial trauma were excluded from the study.

Health care workers used a spoken script to explain the study and give eligible patients the opportunity to decline. Any patient who had all four samples collected is considered as having willingly participated in the study as they allowed the sample collection and the use of the data produced from the sample. This study protocol was deemed to be an operational project by the Office of Human Research Affairs at UnitedHealth Group.

Participants were provided instructions and asked to self-collect tongue, nasal, and MT samples, in that order (see Supplement). Tongue samples were collected with a nylon flocked swab (Copan FLOQSwab 502CS01) via the following steps: 1) Extending the tongue, and 2) firmly but gently brushing the swab along the length of the anterior 2/3 of dorsum of the tongue for 10 seconds. Nasal samples were collected with a foam swab (Puritan 25-1506 1PF100) via the following steps: 1) gently inserting the swab in the vertical position into one nasal passage until there is gentle resistance, 2) leaving the swab in place for 10-15 seconds, rotating the swab, and 3) repeating the procedure on the other side with the same swab. MT samples were collected with a nylon flocked swab (MDL NasoSwab A362CS02.MDL) via the following steps: 1) inserting the swab in the horizontal position until gentile resistance was met, 2) leaving the swab in for 10-15 seconds on each side, rotating the swab, 3) repeating in the other nostril with the same swab. After patient sampling was completed, NP samples were collected by a health care worker using a polyester tipped swab on a skinny wire (Puritan 25-800-2PDBG) via the following steps: 1) pass the swab along the floor of the nose until meeting gentle resistance as the swab touches the posterior pharynx, in the nostril corresponding to the patient’s dominant hand, and 2) rotate the swab several times and withdraw the swab.

All samples were stored in viral transport media and refrigerated at 4°C before shipping on ice packs to a reference laboratory for rRT-PCR testing (Quest Diagnostics, San Juan Capistrano, CA). Patient results were transmitted back to the clinical practice via the standard lab information system and electronic health record protocol. Additionally, cycle threshold (Ct) values for all samples that tested positive for SARS-CoV-2 were reported back to the clinical sites. A higher Ct value corresponds to a lower viral load.

### Statistical Analysis

The study was powered to a one-sided, one-sample test of proportions with a continuity correction to determine whether the percentage of patients with a positive result on the NP test that were also positive for a patient-collected test was significantly greater than 90%, assuming the true sensitivity is 98%. Forty-eight positive NP test results are needed for 80% power at 0.05 significance. Based on recent clinical experience in these centers, we assumed a 9% prevalence of COVID-19 among symptomatic people visiting these five ambulatory centers, resulting in a total sample size of 533 patients to observe 48 positive results. Three separate analyses were performed: one comparing tongue samples to NP samples, a second comparing nasal samples to NP samples, and a third comparing MT samples to NP samples; all used health care worker-collected NP samples are the comparator. Samples included in the final analysis had rRT-PCR results returned for both samples in question (i.e. NP and one patient-collected sample) at the time of data freeze. All statistical analysis was performed using R version 3.6.1^16^.

## Results

We enrolled patients aged 15 months to 94 years old presenting with symptoms indicative of an upper respiratory infection, visiting one of five ambulatory clinical sites in the Puget Sound metropolitan area over five days (March 16 to March 21, 2020). 501 patients had a result for both the tongue and NP samples, 498 had a result for the nasal and NP samples, and 504 had a result for both the MT and NP samples.

Table 1 summarizes the positivity rate in each of the three analysis populations broken out by demographics and self-reported symptoms. Using the NP results, patients had overall positivity rates of 9.8%, 10.0%, and 10.3% for SARS CoV-2 among patients who also returned a tongue, nasal, and MT result, respectively.

**Table 1:**
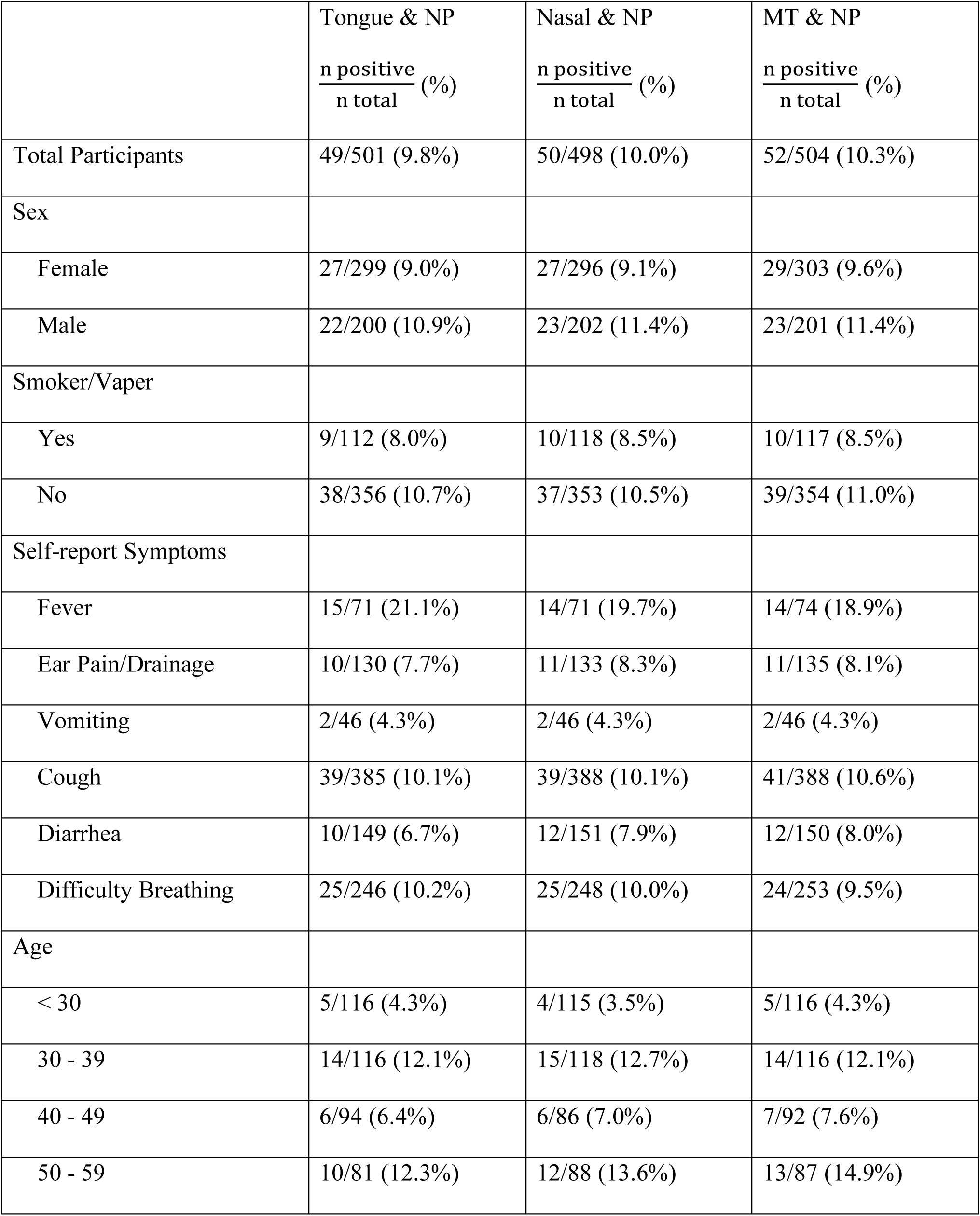
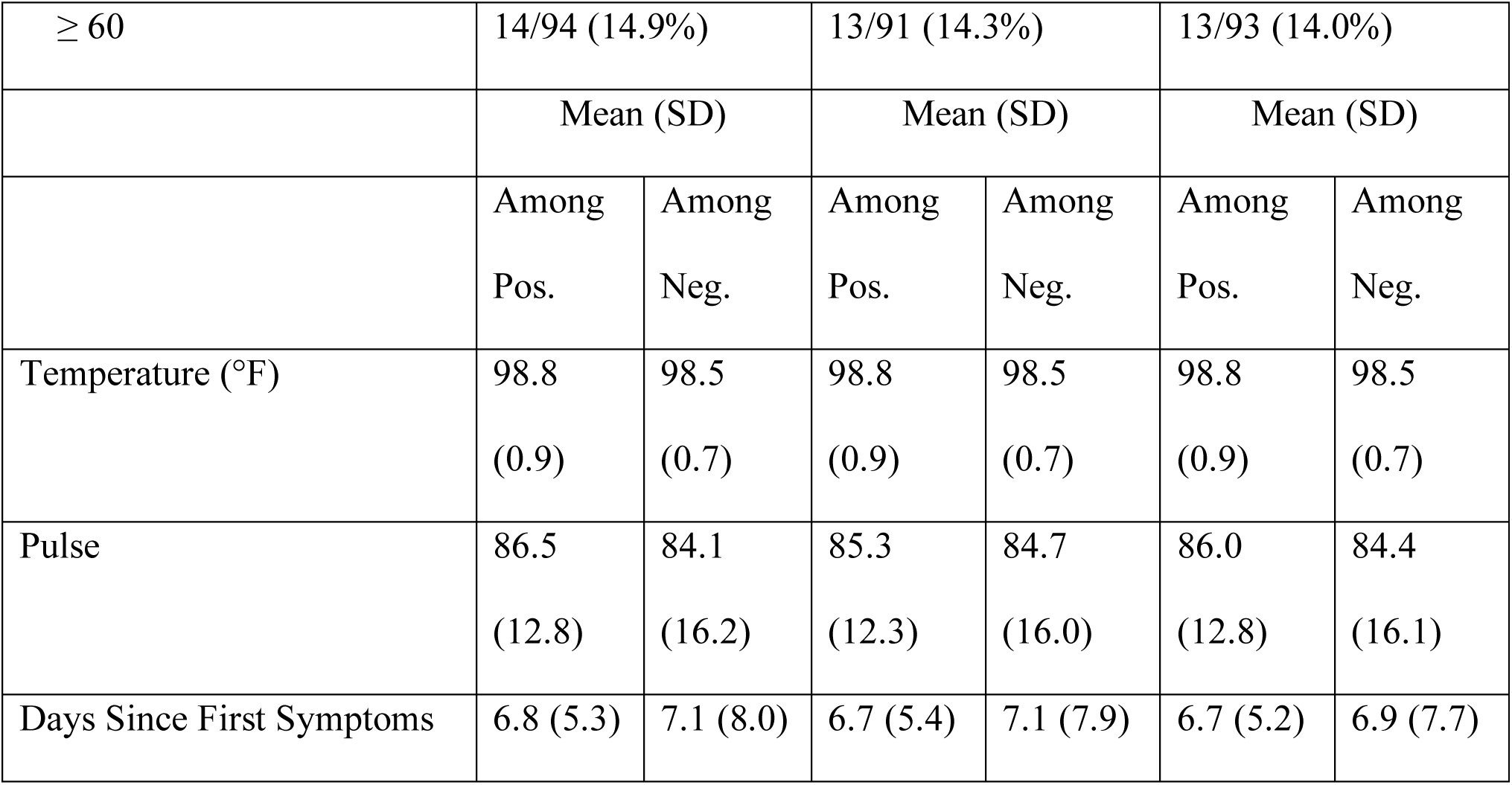
Demographics and self-reported clinical symptoms.

| | Tongue & NP<br>$\frac{n \text{ positive}}{n \text{ total}}$ (%) | Nasal & NP<br>$\frac{n \text{ positive}}{n \text{ total}}$ (%) | MT & NP<br>$\frac{n \text{ positive}}{n \text{ total}}$ (%) |
| --- | --- | --- | --- |
| Total Participants | 49/501 (9.8%) | 50/498 (10.0%) | 52/504 (10.3%) |
| Sex |  |  |  |
| Female | 27/299 (9.0%) | 27/296 (9.1%) | 29/303 (9.6%) |
| Male | 22/200 (10.9%) | 23/202 (11.4%) | 23/201 (11.4%) |
| Smoker/Vaper |  |  |  |
| Yes | 9/112 (8.0%) | 10/118 (8.5%) | 10/117 (8.5%) |
| No | 38/356 (10.7%) | 37/353 (10.5%) | 39/354 (11.0%) |
| Self-report Symptoms |  |  |  |
| Fever | 15/71 (21.1%) | 14/71 (19.7%) | 14/74 (18.9%) |
| Ear Pain/Drainage | 10/130 (7.7%) | 11/133 (8.3%) | 11/135 (8.1%) |
| Vomiting | 2/46 (4.3%) | 2/46 (4.3%) | 2/46 (4.3%) |
| Cough | 39/385 (10.1%) | 39/388 (10.1%) | 41/388 (10.6%) |
| Diarrhea | 10/149 (6.7%) | 12/151 (7.9%) | 12/150 (8.0%) |
| Difficulty Breathing | 25/246 (10.2%) | 25/248 (10.0%) | 24/253 (9.5%) |
| Age |  |  |  |
| < 30 | 5/116 (4.3%) | 4/115 (3.5%) | 5/116 (4.3%) |
| 30 - 39 | 14/116 (12.1%) | 15/118 (12.7%) | 14/116 (12.1%) |
| 40 - 49 | 6/94 (6.4%) | 6/86 (7.0%) | 7/92 (7.6%) |
| 50 - 59 | 10/81 (12.3%) | 12/88 (13.6%) | 13/87 (14.9%) |

|  |  |  |  |  |  |  |
| --- | --- | --- | --- | --- | --- | --- |
| ≥ 60 | 14/94 (14.9%) |  | 13/91 (14.3%) |  | 13/93 (14.0%) |  |
|  | Mean (SD) |  | Mean (SD) |  | Mean (SD) |  |
|  | Among<br>Pos. | Among<br>Neg. | Among<br>Pos. | Among<br>Neg. | Among<br>Pos. | Among<br>Neg. |
| Temperature (°F) | 98.8<br>(0.9) | 98.5<br>(0.7) | 98.8<br>(0.9) | 98.5<br>(0.7) | 98.8<br>(0.9) | 98.5<br>(0.7) |
| Pulse | 86.5<br>(12.8) | 84.1<br>(16.2) | 85.3<br>(12.3) | 84.7<br>(16.0) | 86.0<br>(12.8) | 84.4<br>(16.1) |
| Days Since First Symptoms | 6.8 (5.3) | 7.1 (8.0) | 6.7 (5.4) | 7.1 (7.9) | 6.7 (5.2) | 6.9 (7.7) |

Tables 2, 3, and 4 show 2×2 tables for test results between health care worker - collected NP samples and the patient-collected tongue, nasal, and MT samples, respectively. These tables also provide the estimated sensitivity of the patient-collected samples and one-side 95% confidence intervals. Namely, using health care worker-collected NP samples as the comparator, sensitivity of the patient-collected tongue, nasal, and MT samples were 89.8% (95% CI: 80.2 – 100.0), 94.0% (95% CI 84.6 – 100.0), and 96.2% (95% CI: 87.7 – 100.0), respectively (Tables 2-4).

**Table 2:** A 2×2 table of the test results for all patients who had an NP and a Tongue sample tested.

| Sensitivity (95% CI):<br>89.8% (80.2%, 100.0%) |  | Tongue |  |  |
| --- | --- | --- | --- | --- |
|  |  | Negative | Positive | Total |
| NP | Negative | 450 | 2 | 452 |
|  | Positive | 5 | 44 | 49 |
|  | Total | 455 | 46 | 501 |

**Table 3:** A 2×2 table of the test results for all patients who had an NP and a Nasal sample tested.

| Sensitivity (95% CI):<br>94.0% (84.6%, 100.0%) |  | Nasal |  |  |
| --- | --- | --- | --- | --- |
|  |  | Negative | Positive | Total |
| NP | Negative | 447 | 1 | 448 |
|  | Positive | 3 | 47 | 50 |
|  | Total | 450 | 48 | 498 |

**Table 4:** A 2×2 table of the test results for all patients who had an NP and a MT sample tested.

| Sensitivity (95% CI):<br>96.2% (87.7%, 100%) |  | MT |  |  |
| --- | --- | --- | --- | --- |
|  |  | Negative | Positive | Total |
| NP | Negative | 452 | 0 | 452 |
|  | Positive | 2 | 50 | 52 |
|  | Total | 454 | 50 | 504 |

While the sensitivity of the nasal and MT samples were greater than 90%, none of the patient-sample sensitivities were statistically significant when tested using a one-sided test of proportions (p-values 0.50, 0.24, and 0.11 for tongue, nasal, and MT, respectively). The power calculations, which assumed a true sensitivity of 98%, required 48 positive NP results for each pairwise comparison while the data ultimately showed 49, 50, and 52 NP positives. All three comparisons reached the required sample size, but the observed effect sizes was less than assumed for the power analysis (89.8%, 94.0%, and 95.8% for tongue, nasal, and MT respectively vs 98.0% assumed for the power analysis). Despite this drawback, the estimated sensitivities for nasal and MT samples exceeded 90%. To our knowledge, this study represents the largest available sample directly comparing patient-collected tongue, nasal, and MT samples to health care worker-collected NP samples for COVID-19.

Ct values calculated by the rRT-PCR analysis demonstrated Pearson correlation coefficients of 0.48, 0.78, and 0.86 between the positive NP results the positive tongue, nasal, and MT results, respectively. Figure 1 shows plots of the Ct values for the patient collected sites against the NP site, with a linear regression fit super-imposed on the scatterplot.

**Figure 1:**
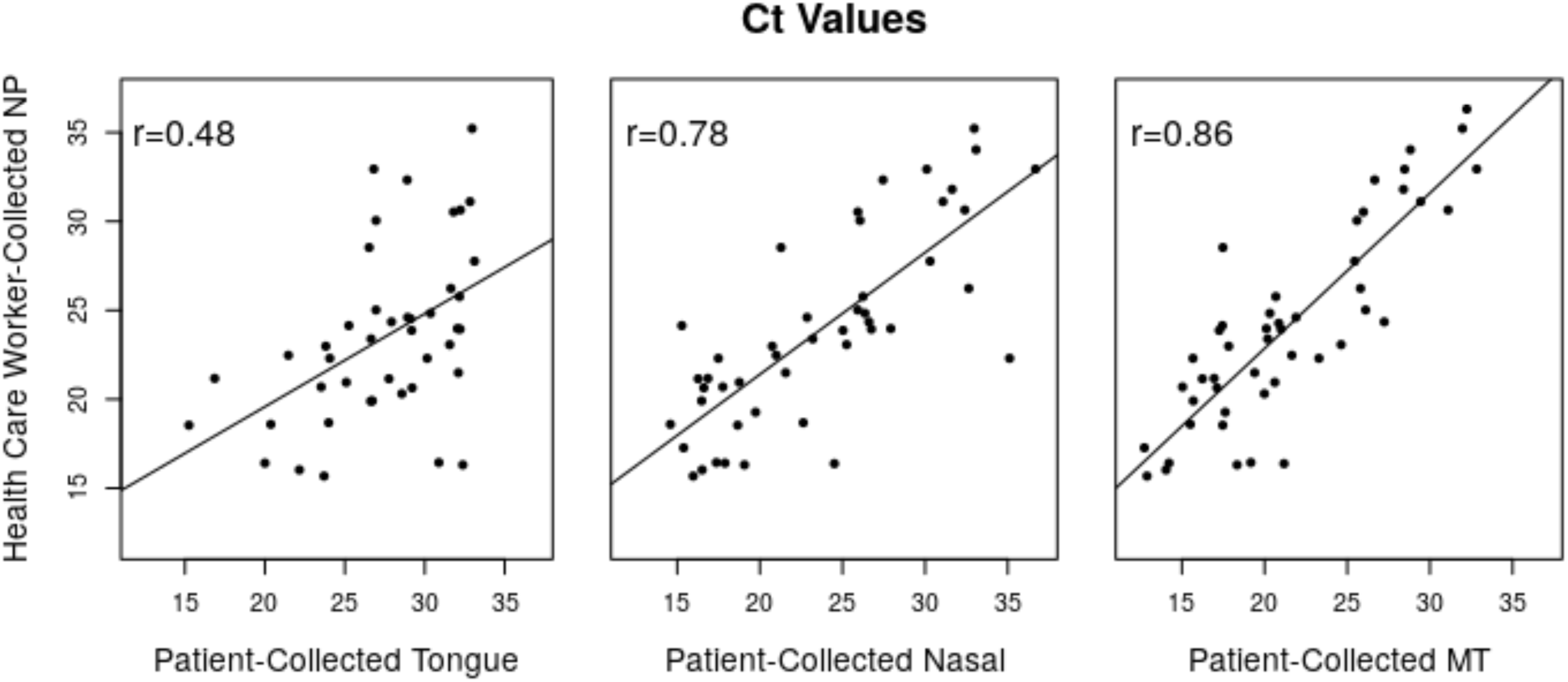
Plots showing the Cycle Threshold (Ct) values of the tongue, nasal, and MT tests against those of the comparator NP test. The correlation coefficient is superimposed on each sub-figure along with a trend line estimated using a simple linear regression. Figure 1a) shows Ct values from the 43 patients that had positive tongue and NP results and available Ct values. Figure 1b) shows Ct values from the 46 patients that had positive nasal and NP results and available Ct values. Figure 1c) shows Ct values from the 48 patients that had positive MT and NP results and available Ct values.

## Discussion

This work demonstrates the clinical utility and equivalency of using patient-collected tongue, nasal, or MT sampling to health care worker-collect NP sampling for diagnosis of COVID-19. Sensitivity of nasal and MT patient collected methods was calculated to be above 90%, and in cohorts of more than 490 patients with respiratory symptoms, patient-collected sampling was feasible in ambulatory practice. The ability to allow patients to self-collect confers a number of benefits to both patient, provider, and system. First, patients are likely to tolerate the alternate collections locations of MT, anterior nares or tongue over NP. NP sampling can cause coughing and sneezing which may be uncomfortable to the patient and increase the risk of aerosol transmission of SARS-CoV2 transmission to health care workers. A patient-collected sample reduces personal protective equipment use, which is currently in short supply. When patients collect their own samples, health care workers can focus on other patients or other parts of the clinical encounter, increasing practice efficiency though optimizing staff utilization.

Other respiratory illnesses have leveraged self-collected samples from locations other than NP. MT collection using a nylon, flocculated swab were found to be equivalent to nurse collected in one study^17^, while self-collected MT swabs were found to be a reliable alternative to health worker collection for influenza A and B virus RT-PCR analysis in another study^18^. Similarly, saliva collected from the tongue has also held promise. In a two-phase study, tongue swabs (two per subject) exhibited a combined sensitivity of 92.8% relative to sputum for tuberculosis detection in adults^12^, and exhibited promise as non-invasive samples for diagnosis of pediatric tuberculosis^13^.

This study has a number of limitations. Samples were collected in five urgent care clinics located in a single region of the US. Our analysis was cross-sectional and limited to single comparisons to NP. With additional analysis and longitudinal data collection, we hope to understand how self-collection of samples from multiple upper respiratory anatomical sites contribute to test performance.

Despite these limitations, we believe that self-collected samples for SARS-CoV-2 testing from sites other than NP is a useful approach during the COVID-19 pandemic.

## Data Availability

A portion of the data, de-identified, has been submitted to FDA for review and public policymaking,.

## ACKNOWLEDGMENTS

The authors acknowledge the contributions of Karen Heichman, Andrew Trister, Daniel Wattendorf, Jessica Lee and Emily Turner from the Bill & Melinda Gates Foundation, Shawna Cooper and Philip Su from Audere, Kris Weigel and Alaina Olson from the University of Washington, John Tamerius from Quidel, Nigel Clarke and James Devlin from Quest Diagnostics, Franklin Cockerill (consultant to Quest Diagnostics), Hunter McCawley and Jiameng Wang from UnitedHealth Group, and Garrett Galbreath, the health care workers, and staff from The Everett Clinic. GA Cangelosi and RC Wood were supported by the Bill & Melinda Gates Foundation.

